# Linguistic Features from Paragraph Recall are Markers of Cognitive Impairment

**DOI:** 10.64898/2025.12.18.25342497

**Authors:** Seho Park, Nicole Roth, Megan Barker, Sanford Auerbach, Thomas T. Perls, Stephanie Cosentino, Rhoda Au, David J. Libon, Paola Sebastiani, Stacy L. Andersen

**Author notes:** Corresponding author: Stacy L. Andersen.

## Abstract

**Objective:** Cognitive impairment is associated with language changes that may be elicited from verbal responses during neuropsychological assessments that are not captured in traditional scoring. The current study investigated the utility of a linguistic analysis of paragraph recall responses for differentiating participants with and without cognitive impairment.

**Methods:** Digital voice recordings of Logical Memory (LM) were available from 598 participants from the Long Life Family Study with normal cognition and 112 with cognitive impairment. Linguistic polyfeature scores for immediate (PFS-IR) and delayed recall (PFS-DR) were created from a weighted sum of features associated with cognitive impairment. Logistic regression models assessed the predictive value of each PFS and demographics for classifying cognitive impairment. Repeated measures models with Generalized Estimating Equations assessed whether PFSs predict decline on a cognitive screener.

**Results:** Both immediate and delayed PFSs were significantly associated with cognitive status (PFS-IR β = 0.05, *p<*.001; PFS-DR β = 0.07, *p<*.001). A classifier with PFS-DR and demographics closely approximated the accuracy of the traditional LM score and demographics (AUC-PR = 0.81 vs 0.84, respectively). A higher PFS-DR was also associated with greater cognitive decline over an average of 5 years of follow-up (β = -0.08, *p<*.001).

**Conclusion:** Quantification of linguistic features from paragraph recall using a linguistic PFS provides sufficient information for detecting cognitive impairment and predicting incident cognitive decline. The linguistic PFS has the potential to be integrated into automated testing, recording, and scoring pipelines allowing for the implementation of sensitive neuropsychological assessments in broader clinical and research settings.

## Background

Deterioration of language performance is frequently observed in patients with cognitive impairment and dementia, and can include semantic impairment, decreases in fluency, and low content density (Fraser et al., 2016; Kempler & Goral, 2008; Taler & Phillips, 2008). However, the traditional scoring of many neuropsychological tests, particularly those that are not classically considered to be language tests, may not capture these changes. Moreover, evidence suggests that disturbances in language can manifest even at the stage of mild cognitive impairment (MCI), preceding more overt signs of dementia (Boschi et al., 2017; Verma & Howard, 2012). By leveraging automated linguistic analysis, these potential early markers can be explored with great efficiency, thereby offering a promising approach for early diagnosis and intervention in dementia.

Recent advances in computational analysis have opened new avenues for detecting speech and language features from voice recordings of neuropsychological assessments. Unlike natural speech, cognitively demanding tasks that require engagement of multiple cognitive functions, such as episodic memory, attention, and executive function, may be more susceptible to impairment in response to early neuropathological changes than the processes typically involved during everyday conversations. For example, a recent study using acoustic features from a delayed paragraph recall task with a high memory load showed better classification accuracy for cognitive status than acoustic features collected from tasks with low to moderate memory loads, such as natural speech and repetition tasks (M. Bae et al., 2023). More broadly, analyses of voice recordings of full neuropsychological examinations from the Framingham Heart Study using various natural language processing methods and acoustic features were able to classify participants by cognitive status with high accuracy (Amini et al., 2023; Tavabi et al., 2022). Other research has focused on specific cognitive tasks that were designed to elicit spontaneous speech, such as picture description tasks. Several studies have shown that linguistic markers, such as semantic content, speech efficiency, and frequency of use of nouns and verbs on the Cookie Theft (CT) task of the Boston Diagnostic Aphasia Examination were able to differentiate between individuals with normal cognition and those with dementia due to Alzheimer’s disease (AD) (Ahmed et al., 2013; Fraser et al., 2016; Kavé & Goral, 2016). Moreover, a longitudinal study found that the presence of altered grammatical structures, repetitiveness, and misspellings among responses from cognitively healthy adults was associated with incident AD, suggesting that these language features may be preclinical markers of decline (Eyigoz et al., 2020).

The Logical Memory (LM) subtest of the Wechsler Memory Scale is a widely used paragraph recall test designed to measure episodic memory, which is commonly affected in normal aging and more substantially in cognitive impairment due to AD and related dementias (Tromp et al., 2015). LM performance actually has one of the strongest predictive values among neuropsychological assessments for future conversion to dementia (Silva et al., 2013). Although this test is primarily described as an episodic memory task, cognitive tests are not domain-specific, often drawing on multiple cognitive processes. For example, the verbal response format of LM also draws on language function, allowing for analysis of linguistic features of a participant’s response. Traditional scoring of LM relies on the total number of story units that are correctly recalled, yet it does not account for related responses that do not meet the strict criteria of allowable responses or responses that are unrelated to the story. However, participants often make additional verbal responses during recall that are rich in cognitive-related information, such as repeating words or phrases, using semantically similar words, or commenting on their performance (e.g., “I can’t remember”). Unlike the previous analyses performed on picture description tasks, which place greater demands on semantic knowledge (Mueller et al., 2018), analysis of LM responses may reveal different linguistic markers, reflecting both episodic memory and language deficits. While recent analysis of a similar paragraph recall task identified acoustic markers that are associated with cognitive impairment (M. Bae et al., 2023), text-based linguistic features remain under-investigated. Additionally, specific subtypes of MCI have different neuropsychological profiles (i.e., amnestic versus dysexecutive profiles) (Bondi et al., 2014; Pa et al., 2009) and thus are likely to be associated with differences in LM recall performance and language use. Taken together, capturing full verbal responses to LM allows us to extract more comprehensive information from a single test that may be more sensitive for detecting cognitive impairment, differentiating MCI subtypes, and predicting cognitive decline.

This study aim to perform a text-based analysis of verbal paragraph recall responses to identify embedded linguistic features that differentiate between cognitively healthy individuals and those with cognitive impairment. We also sought to determine whether linguistic features of LM responses can predict cognitive change over longitudinal follow up.

## Methods

### Study participants and data collection

The Long Life Family Study (LLFS) is a longitudinal family-based cohort study in the United States and Denmark aimed at determining the genetic, behavioral, and environmental factors related to healthy aging and longevity among 4,953 individuals from 539 families selected for evidence of familial longevity (Wojczynski et al., 2022). To date, participants have completed up to three in-home visits (Visit 1: 2006-2009, Visit 2: 2014-2017, Visit 3: 2020-2024) which included a neuropsychological assessment comprised of the Mini Mental State Examination (MMSE), WMS-R Logical Memory, Number Span, semantic (animal) fluency, and the Digit Symbol Substitution Test across all visits and added the Hopkins Verbal Learning Test – Revised (HVLT-R), Clock Drawing Test, Trail Making Test and phonemic fluency at Visit 2 and thereafter. The neuropsychological assessment was voice-recorded at Visit 2 for a subset of LLFS participants (N=710) from the Boston field site. The recordings of the LM immediate recall and delayed recall responses were manually transcribed and cross-checked by a second transcriber to ensure uniform quality.

A study partner completed the Dementia Questionnaire (Kawas et al., 1994; Silverman et al., 1986) to identify cognitive problems in the participant’s daily life. Participants were monitored longitudinally (annually for participants age 70 years and over and every three years for younger participants) for changes in cognitive status using a modified version of the Telephone Interview for Cognitive Status (TICS-m), a brief 12-item test of global cognitive status with a maximum score of 51 points (Brandt et al., 1988; Welsh et al., 1993). Cases flagged for possible cognitive impairment, based on cognitive testing or informant reports of cognitive concerns, were reviewed in clinical consensus conferences with neurologists and neuropsychologists and assigned a Clinical Dementia Rating (CDR) (Hughes et al., 1982).

For this analysis, participants were classified with normal cognition (NC) if they were not flagged for consensus review or were assigned a CDR score of 0 at Visit 2 during consensus review. Participants were classified with cognitive impairment (CI) if they were assigned a CDR score of 0.5 or greater (i.e., mild cognitive impairment or dementia) at Visit 2 during consensus review. Lower than expected performance (i.e., less than 1.5 standard deviations below the demographically-adjusted normative mean) in episodic memory, executive function, attention, or language were also classified during clinical consensus meetings for participants meeting criteria for mild cognitive impairment (CDR=0.5) when sufficient data were available. Participants with CDR=0.5 and lower than expected performance in episodic memory (i.e., HVLT-R and/or LM) were classified as having an amnestic profile (aMCI). Participants with CDR=0.5 and lower than expected performance in executive function (i.e., phonemic fluency, Trail Making Test B, and/or the Digit Symbol Substitution Test) were classified as having a dysexecutive profile (dysMCI). All participants provided written informed consent and each participating institution’s Institutional Review Board reviewed and approved this project.

### Statistical analyses

#### Linguistic feature extraction and feature selection

Linguistic features were extracted with the 2015 version of Linguistic Inquiry and Word Count (LIWC), which is a text analysis tool developed to quantitatively assess the psychological, emotional, and cognitive characteristics of written or spoken language (Tausczik & Pennebaker, 2010). LIWC compares each word in a sample to internal dictionaries and calculates the percentage of words in 93 pre-determined linguistic dimensions including emotional, cognitive, and structural components. The program was developed to measure aspects of language use that may reflect thoughts and emotions, however, these linguistic features have also been used for detection and classification of dementia in conversational speech as well as in story recall and picture description tasks (Asgari et al., 2017; Boland et al., 2024; Haulcy & Glass, 2021; Weiner & Schultz, 2018).

After generating the 93 LIWC variables, we removed 41 variables with either low frequency (e.g., words related to religion or death) or irrelevance to the LM task (e.g., punctuation) since this was a spoken rather than written response. An additional 6 variables in the immediate recall condition and 10 in delayed recall were excluded due to redundancy (Pearson correlation r > 0.6 or r < -0.6). Five different categories of pronouns were combined into three variables: first person, second person, and third person pronouns, resulting in a final set of 44 immediate and 40 delayed recall variables. Since most variables were skewed, we standardized them with median absolute deviation (MAD), which also created beta estimate weights on the same scale. For variables with zero-inflation (22 in immediate; 20 in delayed recall) we used two types of binary transformation (less than median versus median or greater; 0 versus 1+ for variables with a median value of 0).

A previous study using 68 word categories from LIWC to predict cognitive status had decreased model performance, indicating that some word categories are not related to cognitive function (Asgari et al., 2017). Therefore, we performed additional feature selection to remove variables not associated with cognitive status. We used individual regression models for each linguistic feature adjusted by age, sex, and education to predict cognitive status (NC versus CI) and retained only those with a Bonferroni corrected threshold *p* <0.0011. We estimated the regression coefficients using Generalized Estimating Equations (GEE) with exchangeable correlation to account for possible within-family correlation.

#### Linguistic Polyfeature Scores

Since the effect of each linguistic feature associated with cognitive impairment is small, we developed a novel method of quantifying the features into a linguistic polyfeature score (PFS). This method was derived from the concept of polygenic risk scores which quantify an individual’s genetic susceptibility to a specific trait or disease by aggregating the effects of numerous genetic variants across the genome. Although each variant contributes only a small amount of risk, their combined influence can help estimate the likelihood of developing complex conditions such as heart disease, diabetes, or AD (H. Bae et al., 2025; Wray et al., 2008). For the linguistic PFS we combined the individual LIWC metrics from the immediate recall condition that were significantly associated with cognitive impairment into a single weighted sum score called an immediate recall PFS (PFS-IR). Each feature (median absolute deviation standardized or dichotomized) was weighted based on the beta estimate and adjusted for the standard error in the GEE model. Similarly, the individual LIWC features from delayed recall that were associated with cognitive impairment were combined into a delayed recall PFS (PFS-DR).

#### Cross-sectional Analyses

To estimate the association between the linguistic PFS-IR and PFS-DR and cognitive impairment (i.e., NC versus CI), we used logistic regression models for each linguistic PFS adjusted by age, sex, and education to predict cognitive status and estimated the regression coefficients using GEE with exchangeable correlation to account for possible within-family correlation.

To assess the discriminative power of the linguistic PFSs independently and in comparison to traditional cognitive test scores, we built the following five logistic regression models using GEE with exchangeable correlation for within family relatedness and cross-validation to distinguish between CI and NC: 1) PFS-IR, 2) PFS-DR, 3) PFS-IR and PFS-DR, 4) traditional LM delayed recall score, and 5) MMSE score. Each model was adjusted for age, sex, and education. Random sampling was used to balance class distribution by cognitive status (CI versus NC), after which the models were trained on 67% of the data and then tested on the remaining 33%. Due to the imbalanced sample, precision-recall curves, rather than receiver operating characteristic curves, of each classifier were plotted to represent the true positive rate in relation to positive predictive value. For each classifier, we compared evaluation metrics including positive predictive value, negative predictive value, Matthews Correlation Coefficient (MCC), and F1. MCC serves as a holistic complement to traditional metrics for evaluating binary classification quality incorporating critical confusion matrix measures to offer a balanced assessment of model performance and is especially useful for imbalanced classes (Chicco & Jurman, 2023; Richter-Laskowska et al., 2025). The F1 score is the harmonic mean of precision and recall and represents the balance between these two metrics (Rainio et al., 2024). To compare the precision-recall performance of the classifiers, we estimated the 95% confidence interval using bootstrapping with 2,000 resamples. R version 4.3.2 was used for all cross-sectional analyses and the ‘geepack’ package was used for regression models with GEE.

#### Longitudinal analysis

We assessed whether linguistic PFS scores predict change in TICS-m scores over follow-up. For model selection, we used iterative stepwise selection with a significance level of 0.1 (HPGENSELECT procedure in SAS) to determine whether PFS scores are associated with TICS-m. The saturated model included the following covariates: PFS-IR, PFS-DR, age, sex, education, time (months following Visit 2), and an interaction term between PFS-DR and time. Following iterative stepwise selection, we implemented a repeated measures model with GEE to identify the association between PFS and TICS-m over time. In the next two models, we added APOE genotypes to control for the genetic risk conferred by APOE ɛ2 and ɛ4 alleles. APOE alleles were based on the single nucleotide polymorphisms (SNPs) rs7412 and rs429358, which were genotyped using real-time PCR and classified as follows: ɛ2 (rs7412 = T; rs429358 = T), ɛ3 (rs7412 = C; rs429358 = T), and ɛ4 (rs7412 = C; rs429358 = C). We then used binary categorization for the ε2 (0 = ε3ε3, 1 = ε2ε2 or ε2ε3) and ε4 genotypes (0 = ε3ε3, 1 = ε3ε4 or ε4ε4). Participants with the ε2/ε4 genotype were excluded. In the fourth model, the traditional LM delayed score was added as a covariate. Finally, the analysis was repeated using a sample restricted to participants without MCI and dementia at Visit 2 to ensure that results were not driven by participants with prevalent cognitive impairment who would be expected to show more decline after Visit 2 and to understand the predictive power of the linguistic PFS in a cognitively healthy sample. All models were repeated measures models with GEE. Longitudinal analyses were conducted in SAS version 9.4.

#### Secondary analyses

Logistic regression models adjusted by age, sex, and education were used to determine whether each LIWC feature was associated with aMCI in reference to NC at a Bonferroni-corrected threshold of *p*<0.001. We estimated the regression coefficients using GEE to account for possible within-family correlation. Similar logistic regression models with GEE were used to determine the LIWC features that were associated with dysMCI in reference to NC. Participants with both amnestic and executive impairments were included in both analyses. R version 4.3.2 was used for secondary analyses and the ‘geepack’ package was used for regression models with GEE.

## Results

Participant characteristics are summarized in Table 1. Of the 710 participants included in the analysis (mean age = 72.3 ± 11.0 years, 53.5% female), 598 participants had NC and 112 had CI. On average, those in CI group were older, were less likely to have a bachelor’s or higher degree and had lower MMSE and LM scores than the NC group.

**Table 1:** Participant characteristics.

| **Variables** | **Normal**<br>**Cognition**<br>**(NC)** | **Cognitive**<br>**Impairment**<br>**(CI)** | **p value** | **Amnestic**<br>**MCI** | **Dysexecutive**<br>**MCI** |
| --- | --- | --- | --- | --- | --- |
| Participants, N (%) | 598 (84%) | 112 (16%) | - | 24 (4%) | 46 (8%) |
| Age, years, mean (SD) | 69.4 (8.4) | 87.5 ± 11.1 | < .001 | 80.4 (11.3) | 85.2 (10.3) |
| Female, N (%) | 322 (54%) | 55 (52%) | .800 | 10 (42%) | 22 (49%) |
| Education, college degree, N (%) | 425 (71%) | 53 (47%) | < .001 | 17 (71%) | 24 (53%) |
| MMSE, mean (SD) | 29.1 (1.2) | 26.6 (2.9) | < .001 | 26.4 (2.5) | 26.8 (2.1) |
| Logical Memory Immediate, mean (SD) | 14.4 (3.3) | 8.9 (4.6) | < .001 | 8.8 (4.1) | 9.9 (3.6) |
| Logical Memory Delayed, mean (SD) | 13.3 (3.6) | 6.3 (4.4) | < .001 | 5.0 (3.2) | 7.5 (3.9) |

*Note:* The p values represent comparisons between the group with normal cognition and the group with cognitive impairment, NC = normal cognition, CI = cognitive impairment, MCI = mild cognitive impairment, MMSE = Mini-Mental State Examination, SD = standard deviation.

Our initial regression models identified eight features in immediate recall responses and seven features in delayed recall responses that were associated with cognitive impairment as shown in Table 2 (see Supplementary Tables 1 and 2 for the results of all LIWC variables). During immediate recall, the CI group had a higher proportion of negations, words related to cognitive processes (see Table 2 for examples), and words indicating a time orientation in the present than the NC group (all *p<*.001, Table 2). In contrast, some features were less common among participants with CI such as prepositions and words related to semantic categories of biological processes, work, space, and leisure. During delayed recall, the CI group again had a higher proportion of negations and a lower proportion of words related to biological processes and work in their responses in comparison to the NC group. Unique to the delayed recall condition, the CI group had a higher use of first-person pronouns and a lower use of words in the past tense and words related to numbers and drives or words related to motivations in their responses compared to the NC group. Figure 1 presents a Venn diagram summarizing the unique and overlapping features from immediate and delayed recall responses.

**Figure 1.**
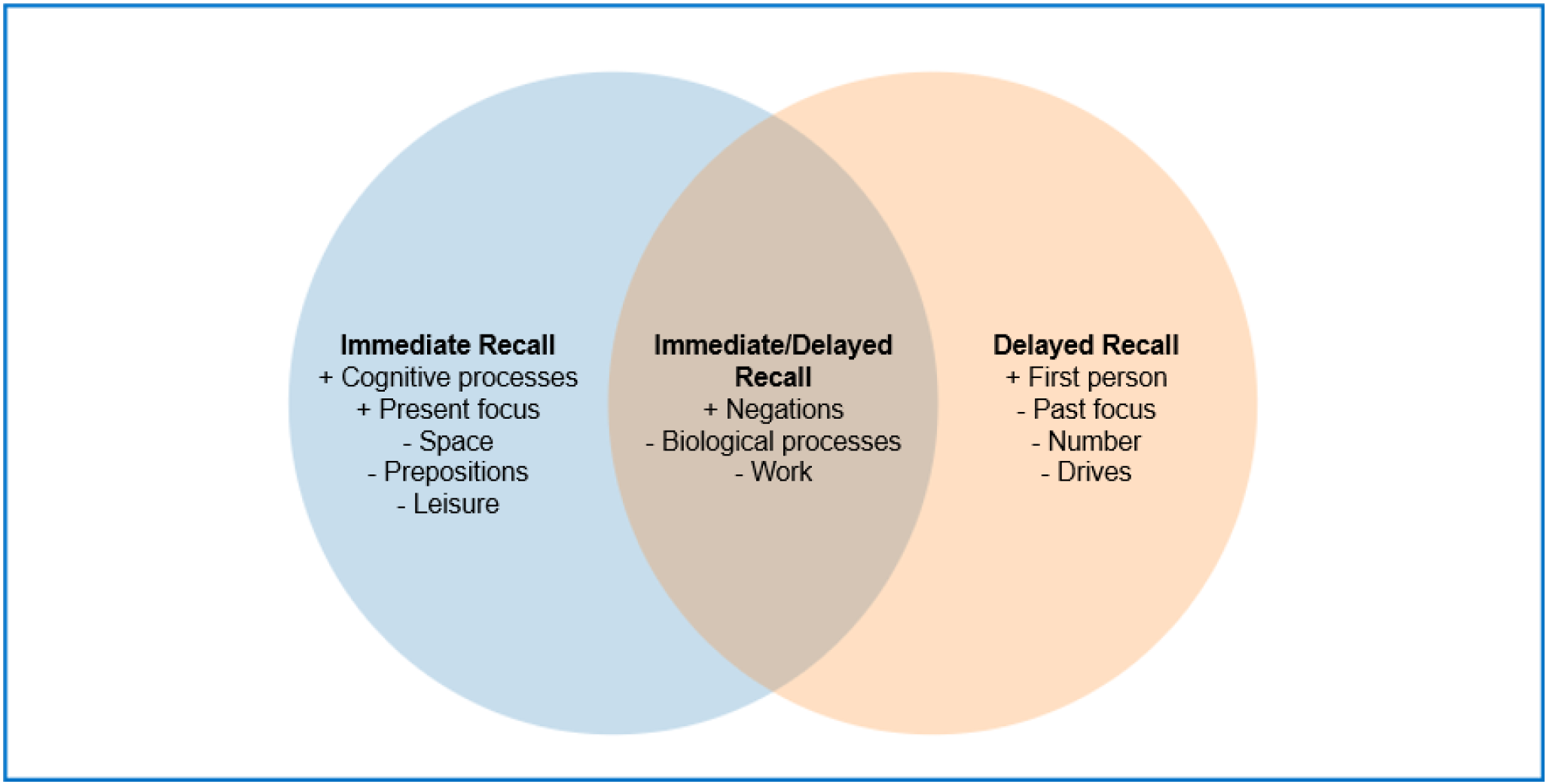
Venn diagram of linguistic features that are associated with cognitive impairment. *Note*: ‘+’ indicates a positive association and ‘-’ indicates a negative association with the outcome of cognitive impairment.

**Table 2:** Linguistic features associated with cognitive impairment.

| LIWC feature | Word Category | Examples | Immediate |  |  | Delayed |  |  |
| --- | --- | --- | --- | --- | --- | --- | --- | --- |
|  |  |  | Beta Estimates | Standard Error | p value | Beta Estimates | Standard Error | p value |
| Negations | Affective Processes | no, not, never | 0.34 | 0.10 | < .001 | 0.64 | 0.11 | < .001 |
| Biological Processes | Biological Processes | cook, cafeteria, eaten | -0.55 | 0.13 | < .001 | -0.58 | 0.15 | < .001 |
| Work | Personal Concerns | job, majors, goal | -0.69 | 0.14 | < .001 | -0.69 | 0.19 | < .001 |
| Cognitive Processes | Cognitive Processes | cause, know, ought | 0.58 | 0.14 | < .001 | - | - | - |
| Present Focus | Time Orientations | today, is, now | 0.37 | 0.11 | < .001 | - | - | - |
| Space | Relativity | down, in, thin | -0.64 | 0.17 | < .001 | - | - | - |
| Prepositions | Linguistic Dimensions | to, with, above | -0.72 | 0.15 | < .001 | - | - | - |
| Leisure | Personal Concerns | cook, chat, movie | -1.32 | 0.28 | < .001 | - | - | - |
| First Person | Personal Pronouns | I, me, mine | - | - | - | 0.53 | 0.11 | < .001 |
| Past focus | Time Orientations | had, did, held, eaten | - | - | - | -0.46 | 0.13 | < .001 |
| Number | Other Grammar | Fifty-six, four, two | - | - | - | -0.51 | 0.14 | < .001 |
| Drives | Drives | took, children | - | - | - | -0.56 | 0.17 | < .001 |

*Note:* Bonferroni corrected threshold $p < 0.0011$ . LIWC = Linguistic Inquiry and Word Count.

PFS-Immediate Recall (PFS-IR) and PFS-Delayed Recall (PFS-DR) scores were created for each participant using the weighted sum of the features associated with cognitive impairment from immediate recall and delayed recall, respectively. The distributions of the PFS-IR scores (median: -4.5, range: -50.8 to 121.3) and PFS-DR scores (median: -0.9, range: -38.6 to 176.2) are shown in Supplementary Figure 1. Linguistic PFS scores had a moderate negative correlation with traditional scores of LM for both immediate and delayed recall (Spearman correlation coefficient, *ρ* = -0.531, *p<*.001; *ρ* = -0.527, *p<*.001, respectively, Supplementary Figure 2), indicating that the PFS, in part, captures different information than traditional scoring. Supplementary Figure 3 shows an example of two participants who both scored 7 on delayed recall using traditional scoring, but their PFS scores differed by more than 50 points.

Both PFSs were highly associated with cognitive status (PFS-IR β = 0.05, *p<*.001; PFS-DR β = 0.07, *p<*.001, Supplementary Table 3), with a higher linguistic PFS being associated with cognitive impairment. As shown in Figure 2, classifiers with either the immediate or delayed PFS with demographic information (PFS-IR: AUC-PR = 0.77, PFS-DR: AUC-PR = 0.81) were more robust than a classifier with only demographic information (AUC-PR = 0.70). In comparison to the full model with both PFS-IR and PFS-DR, the more parsimonious classifier with only PFS-DR and demographics performed slightly better, however, this was not statistically significant (AUC-PR = 0.80 and 0.81, respectively, bootstrapped difference: M = 0.001, 95% CI [−0.001, 0.03] *p* = 0.544). This PFS-DR and demographics classifier also performed similarly to a classifier with traditional LM scores along with demographics (AUC-PR = 0.84).

**Figure 2.**
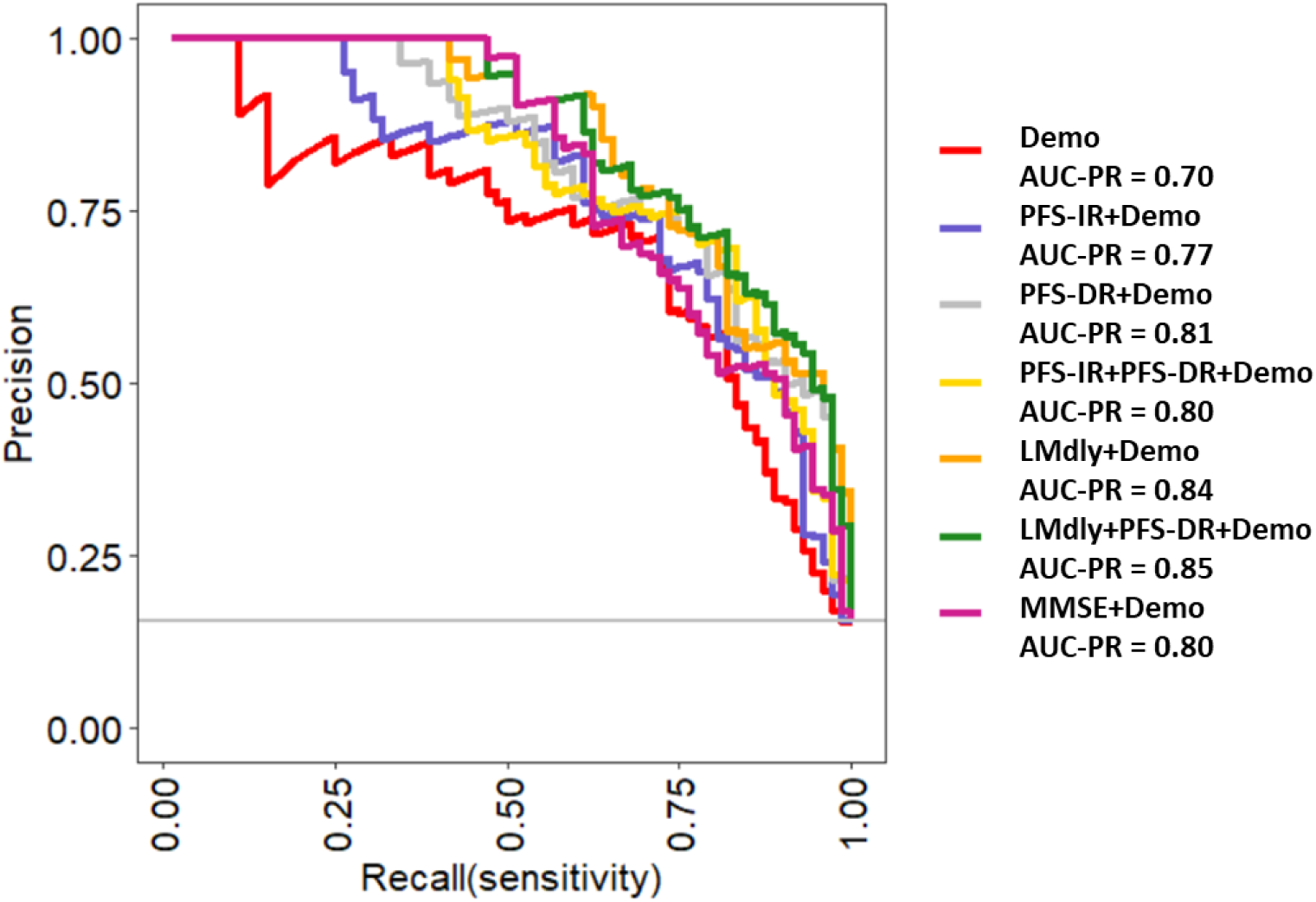
Precision-recall curves for classification of cognitive status. *Note*: Demo = demographics, PFS-IR = immediate recall linguistic polyfeature score, PFS-DR = delayed recall linguistic polyfeature score, Lmdly = traditional logical memory delayed score, MMSE = Mini-Mental State Examination score.

When comparing the performance of PFS-DR and demographics with a cognitive screener (i.e., MMSE) and demographics for detecting cognitive impairment, we found similar performance (AUC-PR = 0.81 and 0.80 respectively, bootstrapped difference: M = 0.006, 95% CI [−0.010, 0.024] *p* = 0.462). Optimizing for the thresholds with the highest Matthews Correlation Coefficient (MCC) the PFS-DR model overall had greater predictive value (MCC = 0.64, F1 = 0.67, Positive Predictive Value [PPV] = 0.83, Negative Predictive Value [NPV] = 0.93) than the model with MMSE (MCC = 0.61, F1 = 0.67, PPV = 0.63, NPV = 0.95). In comparing these two models, we observed the greatest difference was in PPV, and so we estimated 95% CI using bootstrapping with 2,000 resamples and the PPV difference was borderline significant (bootstrapped difference in PPV: M = 0.145, 95% CI [-0.027, 0.314], *p* = 0.110).

A subset of 447 participants completed at least two TICS-m after their digitally-recorded in-person visit with an average follow-up time of 5 years (mean = 61 months, standard deviation [SD] = 17 months, range = 6 - 99 months). Table 3 shows the results for the longitudinal analysis in which PFS-DR had a significant negative association with TICS-m scores adjusting for age, sex, education, PFS-IR, the traditional LM score, and months of follow-up (β = -0.05, *p<*.001). Since the most extreme linguistic PFS scores are likely to be the most informative for identifying preclinical cognitive impairment, a 30-point higher PFS-DR score (i.e., approximately 2 SD in this sample) translates to an almost 3 point difference in TICS-m score over 5 years. Next, we added APOE ε2 and ε4 genotype to the base model, however, the results were largely unchanged (Supplementary Tables 4 and 5). Finally, the base model was applied to a restricted sample omitting participants with cognitive impairment at baseline (n = 63) who are more likely to experience cognitive decline, leaving a sample of 384 participants with NC. PFS-DR remained a significant predictor of TICS-m scores over follow-up (β = -0.02, *p* = 0.041, Supplementary Table 6).

**Table 3:** Prediction of TICS-m scores including traditional logical memory scores over 5 years of follow-up.

| Variables | Estimates | Standard Error | p value |
| --- | --- | --- | --- |
| Age | -0.24 | 0.02 | < .001 |
| Sex (Male) | -0.69 | 0.31 | .025 |
| Education (Advanced degree) | 1.32 | 0.38 | < .001 |
| Education (Bachelor's degree) | 1.07 | 0.36 | .003 |
| Education (High School or less) | - | - | - |
| PFS-IR | 0.02 | 0.01 | .143 |
| PFS-DR | -0.05 | 0.02 | < .001 |
| Months after Logical Memory | -0.02 | 0.003 | < .001 |
| Logical Memory Score | 0.30 | 0.05 | < .001 |

*Note:* TICS-m = Modified Telephone Interview for Cognitive Status, PFS-IR = immediate recall linguistic polyfeature score, PFS-DR = delayed recall linguistic polyfeature score.

### Secondary analyses

Of the participants with cognitive impairment, 24 had an amnestic profile and 46 had a dysexecutive profile. As shown in Table 4, participants with aMCI had a higher percentage of function words and a lower percentage of leisure-related words in their immediate recall responses compared to participants with NC. Among delayed recall features, participants with aMCI had a higher percentage of negations, present-focused words, first-person pronouns, authentic language, and words related to cognitive process in their delayed recall responses. In contrast, they had a lower percentage of clout-related language (i.e., a summary measure of confidence in responses) and work-related words compared to the NC group.

**Table 4:** Linguistic features associated with amnestic mild cognitive impairment.

| LIWC Variables | Word Category | Examples | Beta Estimates | Standard Error | p value |
| --- | --- | --- | --- | --- | --- |
| Immediate Recall Feature |  |  |  |  |  |
| Function words | Linguistic Dimensions | it, to, no, very | 0.80 | 0.23 | < .001 |
| Leisure | Personal Concerns | cook, chat, movie | -1.42 | 0.42 | < .001 |
| Delayed Recall Feature |  |  |  |  |  |
| Negations | Affective Processes | no, not, never | 1.17 | 0.18 | < .001 |
| Present Focus | Time Orientations | today, is, now | 1.06 | 0.18 | < .001 |
| First Person | Personal Pronouns | I, me, mine | 1.03 | 0.30 | < .001 |
| Authentic | Summary Language Variable | personal content | 0.75 | 0.17 | < .001 |
| Cognitive Processes | Cognitive Processes | cause, know, ought | 0.71 | 0.15 | < .001 |
| Clout | Summary Language Variable | confidence in content | -0.81 | 0.15 | < .001 |
| Work | Personal Concerns | job, majors, goal | -1.53 | 0.41 | < .001 |

*Note:* Bonferroni corrected threshold $p < 0.0011$ . LIWC = Linguistic Inquiry and Word Count.

Participants with a dysexecutive profile of MCI had a lower percentage of words related to biological processes and leisure in their immediate recall responses compared to participants with NC as shown in Table 5. In delayed recall, participants with dysMCI had a higher percentage of present-focused words and negations, but a lower percentage of clout-related language and past-focused words, compared to the NC group.

**Table 5:** Linguistic features associated with dysexecutive mild cognitive impairment.

| LIWC Variables | Word Category | Examples | Beta Estimates | Standard Error | p value |
| --- | --- | --- | --- | --- | --- |
| Immediate Recall Features |  |  |  |  |  |
| Biological Processes | Biological Processes | cook, cafeteria, eaten | -0.69 | 0.18 | < .001 |
| Leisure | Personal Concerns | cook, chat, movie | -1.22 | 0.35 | < .001 |
| Delayed Recall Features |  |  |  |  |  |
| Present Focus | Time Orientations | today, is, now | 0.63 | 0.16 | < .001 |
| Negations | Affective Processes | no, not, never | 0.57 | 0.15 | < .001 |
| Clout | Summary Language Variable | confidence in content | -0.52 | 0.14 | < .001 |
| Past Focus | Time Orientations | ago, did, talked | -0.68 | 0.19 | < .001 |

*Note:* Bonferroni corrected threshold $p < 0.0011$ . LIWC = Linguistic Inquiry and Word Count.

## Discussion

We found that linguistic features extracted from a paragraph recall task were able to detect cognitive impairment and predict change in cognition over time. We used a readily available text analysis tool to extract linguistic features from a commonly used test of episodic memory. The most notable features that were associated with cognitive impairment were prepositions, negations, pronouns, and words related to time orientation and the contents of the story. These findings add to the growing body of evidence that linguistic markers can be extracted from tests not necessarily designed to examine language function, and may be meaningful for detecting cognitive impairment (Amini et al., 2023; de la Fuente Garcia et al., 2020; Petti et al., 2020; Tavabi et al., 2022). The current analysis specifically focused on semantic and linguistic features from participants’ verbal responses on a paragraph recall task. Our findings align with prior work showing that both semantic content (e.g., story-related terms) and lexical features (e.g., preposition use) in spoken and written responses are clinically informative for assessing cognitive status and detecting preclinical impairment (Ahmed et al., 2013; Eyigoz et al., 2020; Petti et al., 2020; Taler & Phillips, 2008; Weissenbacher et al., 2016).

### Summary scores of linguistic features detect cognitive impairment and predict cognitive decline

Subtle linguistic features, rather than coarse linguistic measures such as total word count, were associated with cognitive status but had small effect sizes. Thus, the creation of linguistic PFSs (i.e., the combined summary scores of these features) was key to the reliable differentiation of participants with CI from those with NC. The PFS-DR achieved classification accuracy comparable to that of a widely used cognitive screener, the MMSE, and notably outperformed the MMSE in terms of positive predictive value. Moreover, the MMSE is known to suffer from poor sensitivity in early stages of dementia in comparison to verbal episodic memory and language tasks (Sabe et al., 1993). Therefore, our approach could enable the implementation of sensitive and specific neuropsychological assessments into cognitive screening processes, particularly if integrated with an automated pipeline for digital voice capture, auto-transcription, and scoring in a seamless process. These tools can also be readily adapted for other languages (e.g., LIWC has dictionaries in more than 15 different languages), thereby extending their applicability to diverse linguistic populations and potential to enhance cross-cultural research. In our longitudinal analysis, we found that the linguistic PFS predicted future decline on a cognitive screener over 5 years of follow-up. These findings were robust to restriction of the sample to participants with normal cognition at the time of the voice recording and supports the potential value of the linguistic PFS as an indicator of preclinical cognitive changes. Furthermore, our results showed that the linguistic PFS predicted future decline on a cognitive screener even when including the traditional LM score, suggesting that it provides additional information about incident cognitive decline that is not captured by traditional, manual scoring of the LM task.

### Individual linguistic features are associated with cognitive impairment

As mentioned above, coarse linguistic measures, such as total word count, were not associated with cognitive status, as also seen in other studies (Chapin et al., 2022; Kavé & Goral, 2016). One previous study did find a difference in word count (König et al., 2024), however, they used much longer discourse than the logical memory responses in our study, which generally contained less than 60 seconds of content. In our study, a lower proportion of content words, such as those related to work, leisure, and numbers, was also associated with cognitive impairment, which is perhaps expected as these categories are closely related to the content of the stimulus story. This suggests that participants with cognitive impairment conveyed less content information than participants with normal cognition, which aligns with a deficit in episodic memory. Specific to the immediate recall condition, a lower use of prepositions was associated with cognitive impairment. This is consistent with findings from Lindsay and colleagues of “semantically empty language” and reduced use of prepositions during a picture description task among individuals with dementia due to AD (Lindsay et al., 2021), and suggests poorer encoding of the story details among those with CI in our study. Additionally, a greater use of ‘negations’ and words related to ‘cognitive processes’ in participants’ immediate recall responses was associated with an increased likelihood of cognitive impairment in our study. Responses such as “don’t know” or “can’t remember” contribute to these linguistic categories and may reflect an awareness of cognitive deficits, uncertainty in their responses, or a hesitancy to respond. In the delayed recall condition, there was an enrichment of the use of first-person pronouns and a lower use of the past tense among participants with cognitive impairment, which may reflect a shift toward commenting about their performance instead of adhering to the task of providing story details as the story is set in the past tense. This result is partially congruent with a previous study that found a higher proportion of pronouns on immediate, rather than delayed recall, of multiple stories among participants with MCI (Boland et al., 2024). Linguistic features that are unique to each recall condition should be further investigated as they may elucidate specific processes underlying deficits in learning and recall.

These findings show that information-rich responses from participants’ responses hold nuances that are not fully captured by the traditional LM scoring system. Clinicians practicing the "Boston Process Approach" have long appreciated that qualitative features of a person’s process while completing a task, not just their final score, are informative for understanding cognitive function (Ashendorf et al., 2013; Milberg et al., 2009). Some features, such as repetitions of the contents of the story or self-commentary (e.g., “I don’t remember”) are feasible for a clinician to detect, whereas more subtle features (e.g., preposition use) are less so. Digital recording of responses and the NLP techniques used in the current study allow for the systematic quantification of these subtle, process-based features, and also make them accessible to the general clinician.

### Subtypes of cognitive impairment are associated with distinct linguistic profiles

Some linguistic features were similarly altered in those with amnestic MCI or dysexecutive MCI in comparison to cognitively healthy participants. However, amnestic MCI was uniquely associated with a higher proportion of first-person words, cognitive process words, and a higher ‘authentic’ score (i.e., a summary measure of personal content; Tausczik & Pennebaker, 2010) which taken together suggest a shift toward talking about oneself rather than recalling the story. These distinct patterns of word use and linguistic features may aid in the early differentiation of MCI subtypes, however, these results must be interpreted with caution as the amnestic MCI and dysexecutive MCI sample sizes were small.

### Limitations

A limitation of the current study is that the clinical consensus conferences included review of traditional LM scores as part of the determination of cognitive status. As a result, the content of the LM responses, and therefore the LIWC variables, is not completely independent of the outcome variable of cognitive status. This circularity issue is likely to be even more pronounced for the aMCI group in the MCI subtype analysis. Additionally, this study was limited to linguistic features that are coded by the LIWC program. The program was primarily designed to assess psychological constructs, such as positive and negative affect, whereas LM was developed to assess episodic memory. Therefore, LIWC may not capture all relevant linguistic features associated with learning and recall. Additionally, our study may have been underpowered to detect all of the LIWC features that are associated with cognitive impairment due to the small sample size. For example, word count trended toward significance (β = -0.40, *p* = 0.005) but did not meet the Bonferroni-corrected threshold. It must also be noted that the LIWC program does not incorporate the syntactic structure of responses and thus may not accurately capture word meanings that vary depending on context within a sentence. Lastly, this study focused on a text-based, rather than audio-based, analysis of voice recordings. Future studies should integrate additional features from digital voice recordings, such as acoustic and temporal patterns of responses, which have the potential to increase the sensitivity and specificity of determining cognitive status and detecting preclinical cognitive impairment.

In summary, this study identified novel, subtle features of verbal responses on a brief assessment of episodic memory that were informative for detecting cognitive impairment and predicting incident decline. These methods extract additional clinically-useful information from a single cognitive test and may enable automated scoring of paragraph recall tests, which are more sensitive and specific to deficits related to neurodegenerative diseases than cognitive screeners typically used in clinical settings.

## Data Availability

The ELITE portal (https://eliteportal.synapse.org/) provides access to phenotypic and other LLFS data. Additional data produced in the present study are available upon reasonable request to the authors.

## Acknowledgements

We thank the participants of the Long Life Family Study for the generous donation of their time and data and their contribution to knowledge about healthy aging.

