## Supplemental Materials for "Linguistic Features from Paragraph Recall are Markers of Cognitive Impairment"

**Supplementary Table 1**: LIWC features explored for association with cognitive impairment in immediate recall

| Word Category | LIWC features | Estimate | Standard Error | p value |
|---|---|---|---|---|
| Affective Processes | Negations | 0.34 | 0.10 | <.001* |
| | Affective processes[2] | -0.17 | 0.26 | .516 |
| Biological Processes | Biological Processes | -0.55 | 0.13 | <.001* |
| Cognitive Processes | Cognitive Processes | 0.58 | 0.14 | <.001* |
| | Discrepancy[1] | 0.84 | 0.35 | .015 |
| | Insight[2] | 0.56 | 0.25 | .026 |
| | Causation[1] | -0.55 | 0.39 | .159 |
| | Differentiation[2] | 0.46 | 0.33 | .165 |
| | Tentative[1] | 0.23 | 0.28 | .415 |
| | Certainty[1] | 0.04 | 0.27 | .874 |
| Drives | Drives | -0.22 | 0.14 | .113 |
| Informal Language | Nonfluencies[2] | 0.35 | 0.27 | .188 |
| Linguistic Dimensions | Prepositions | -0.72 | 0.15 | <.001* |
| | Article | -0.39 | 0.14 | .007 |
| | Total Function Words | 0.41 | 0.16 | .010 |
| | Impersonal pronouns | 0.29 | 0.14 | .033 |
| | Common Adverbs[2] | 0.47 | 0.29 | .098 |
| | Conjunctions | 0.15 | 0.12 | .207 |
| Other Grammar | Verbs | 0.32 | 0.12 | .006 |
| | Common adjectives[2] | -0.75 | 0.29 | .009 |
| | Number | -0.42 | 0.16 | .009 |
| | Comparison[1] | -0.68 | 0.27 | .013 |
| | Interrogatives[1] | -0.31 | 0.26 | .241 |
| | Quantifiers[1] | 0.16 | 0.28 | .574 |

| | | | | |
|---|---|---|---|---|
| Perceptual Processes | Perceptual Processes[2] | -0.56 | 0.31 | .076 |
| Personal Concerns | Work | -0.67 | 0.14 | <.001* |
| | Leisure[1] | -1.32 | 0.28 | <.001* |
| | Home[1] | -0.59 | 0.3 | .049 |
| | Money | -0.12 | 0.14 | .405 |
| Personal Pronouns | 1st Person Pronoun[2] | 0.48 | 0.29 | .102 |
| | 3rd Person Pronoun | -0.39 | 0.28 | .169 |
| | 2nd Person Pronoun[1] | 0.04 | 0.38 | .921 |
| Relativity | Space | -0.64 | 0.17 | <.001* |
| | Time[2] | -0.46 | 0.28 | .109 |
| | Motion[1] | -0.25 | 0.28 | .364 |
| Social Processes | Female | 0.04 | 0.12 | .723 |
| Summary Language Variable | Clout | -0.34 | 0.11 | .003 |
| | Word Count | -0.40 | 0.14 | .005 |
| | Words per sentence | -0.38 | 0.15 | .011 |
| | Words > 6 letters | -0.24 | 0.14 | .078 |
| | Authentic[2] | 0.05 | 0.29 | .851 |
| Time Orientation | Present Focus | 0.37 | 0.11 | <.001* |
| | Past Focus | -0.13 | 0.10 | .216 |
| | Future Focus[1] | 0.31 | 0.40 | .445 |

*Note:* Bonferroni corrected threshold $p$ <.0011*. [1]Dichotomized into 0 versus 1 or more. [2] Dichotomized into less than median versus median or greater. All other variables were standardized with median absolute deviation.

**Supplementary Table 2**: LIWC features explored for association with cognitive impairment in delayed recall

| Word Categories | LIWC Features | Estimate | Standard Error | p value |
|---|---|---|---|---|
| Affective Processes | Negations | 0.64 | 0.11 | <.001* |
| | Affect[2] | 0.01 | 0.25 | .968 |
| Biological Processes | Biological Processes | -0.58 | 0.15 | <.001* |
| Cognitive Processes | Discrepancy[1] | -0.66 | 0.35 | .061 |
| | Causation[1] | -0.74 | 0.42 | .076 |
| | Certainty[1] | -0.14 | 0.29 | .638 |
| | Differentiation[1] | 0.07 | 0.34 | .829 |
| | Tentative[2] | 0.02 | 0.25 | .928 |
| Drives | Drives | -0.56 | 0.17 | <.001* |
| Informal Language | Nonfluencies[2] | 0.71 | 0.28 | .010 |
| Linguistic Dimensions | Prepositions | -0.57 | 0.18 | .002 |
| | Conjunctions | -0.37 | 0.15 | .012 |
| | Total Function Words | 0.36 | 0.15 | .017 |
| | Impersonal Pronouns | 0.27 | 0.12 | .027 |
| | Common Adverbs[1] | -0.53 | 0.29 | .072 |
| | Common Adjectives[2] | -0.04 | 0.29 | .894 |
| Other Grammar | Number | -0.51 | 0.14 | <.001* |
| | Comparison[1] | -0.43 | 0.3 | .160 |
| | Interrogatives[2] | -0.37 | 0.27 | .172 |
| | Verb | 0.17 | 0.13 | .179 |
| | Quantifiers[2] | 0.12 | 0.28 | .672 |
| Perceptual Processes | Perceptual Processes[1] | -0.67 | 0.27 | .015 |
| Personal Concerns | Work | -0.69 | 0.19 | <.001* |
| | Home[1] | -1.09 | 0.39 | .005 |

| | | | | |
|---|---|---|---|---|
| | Leisure[2] | -0.94 | 0.36 | .009 |
| | Money | -0.44 | 0.17 | .010 |
| Personal Pronouns | 1st Person Pronouns | 0.53 | 0.11 | <.001* |
| | 3rd Person Pronoun | -0.63 | 0.29 | .033 |
| | 2nd Person Pronoun[1] | 0.39 | 0.39 | .315 |
| Relativity | Motion[1] | -0.32 | 0.27 | .239 |
| | Time[2] | -0.28 | 0.28 | .322 |
| | Space | -0.2 | 0.31 | .517 |
| Social Processes | Female | -0.38 | 0.14 | .006 |
| Summary Language Variable | Word Count | -0.54 | 0.18 | .002 |
| | Words per sentence | -0.57 | 0.19 | .003 |
| | Authentic[2] | 0.80 | 0.28 | .004 |
| | Words > 6 letters | -0.25 | 0.14 | .080 |
| | Articles | -0.18 | 0.20 | .380 |
| Time Orientation | Past Focus | -0.46 | 0.13 | <.001* |
| | Future Focus[1] | 0.25 | 0.33 | .446 |

*Note:* Bonferroni corrected threshold $p$ <.0011*. [1]Dichotomized into 0 versus 1 or more. [2] Dichotomized into less than median versus median or greater. All other variables were standardized with median absolute deviation.

**Supplementary Table 3**: Logistic regression models predicting cognitive impairment

| Variables | Estimates | Standard Error | p value |
|---|---|---|---|
| PFS-IR | 0.05 | 0.01 | < .001 |
| Age | 0.14 | 0.01 | < .001 |
| Sex | -0.42 | 0.29 | .141 |
| Education | -0.06 | 0.06 | .307 |

| Variables | Estimates | Standard Error | p value |
|---|---|---|---|
| PFS-DR | 0.07 | 0.01 | < .001 |
| Age | 0.15 | 0.02 | < .001 |
| Sex | -0.35 | 0.31 | .250 |
| Education | -0.06 | 0.06 | .380 |

*Note*: PFS-IR = immediate recall linguistic polyfeature score, PFS-DR = delayed recall linguistic polyfeature score.

**Supplementary Table 4**: Prediction of TICS-m scores including APOE ε2 genotype over 5 years of follow-up

| Variables | Estimates | Standard Error | p value |
|---|---|---|---|
| Age | -0.26 | 0.02 | < .001 |
| Sex (Male) | -1.33 | 0.34 | < .001 |
| Education (Advanced degree) | 1.19 | 0.45 | .008 |
| Education (Bachelor's degree) | 1.04 | 0.43 | .015 |
| Education (High School or less) | - | - | - |
| PFS-IR | -0.01 | 0.01 | .264 |
| PFS-DR | -0.09 | 0.02 | < .001 |
| Months after Logical Memory | -0.02 | 0.004 | < .001 |
| ε2 Genotype [1] | -0.33 | 0.41 | .425 |

*Note*: [1] APOE ε4 was coded as 0 = ε3ε3 genotype and 1 = ε2ε2 or ε2ε3 genotype. Participants with ε2ε4 allele were excluded from the analysis. TICS-m = Modified Telephone Interview for Cognitive Status, PFS-IR = immediate recall polyfeature score, PFS-DR = delayed recall polyfeature score.

**Supplementary Table 5**: Prediction of TICS-m scores including APOE ε4 genotype over 5 years of follow-up

| Variables | Estimates | Standard Error | p value |
|---|---|---|---|
| Age | -0.26 | 0.02 | < .001 |
| Sex (Male) | -1.41 | 0.33 | < .001 |
| Education (Advanced degree) | 1.23 | 0.44 | .005 |
| Education (Bachelor's degree) | 1.20 | 0.42 | .004 |
| Education (High School or less) | - | - | - |
| PFS-IR | -0.0001 | 0.01 | .991 |
| PFS-DR | -0.10 | 0.02 | < .001 |
| Months after Logical Memory | -0.02 | 0.004 | < .001 |
| ε4 Genotype [1] | 0.16 | 0.38 | .674 |

*Note*: [1] APOE ε4 was coded as 0 = ε3ε3 genotype and 1 = ε3ε4 or ε4ε4 genotype. Participants with ε2ε4 allele were excluded from the analysis. TICS-m = Modified Telephone Interview for Cognitive Status, PFS-IR = immediate recall linguistic polyfeature score, PFS-DR = delayed recall linguistic polyfeature score.

**Supplementary Table 6**: Prediction of TICS-m score over 5 years of follow-up among participants with normal cognition at baseline

| **Variables** | **Estimates** | **Standard Error** | **p value** |
|---|---|---|---|
| Age | -0.18 | 0.02 | < .001 |
| Sex (Male) | -1.36 | 0.25 | < .001 |
| Education (Advanced degree) | 1.73 | 0.33 | < .001 |
| Education (Bachelor's degree) | 1.16 | 0.31 | < .001 |
| Education (High School or less) | - | - | - |
| PFS-IR | 0.01 | 0.01 | .171 |
| PFS-DR | -0.02 | 0.01 | .041 |
| Months after Logical Memory | -0.02 | 0.004 | < .001 |

*Note*: TICS-m = Modified Telephone Interview for Cognitive Status, PFS-IR = immediate recall linguistic polyfeature score, PFS-DR = delayed recall linguistic polyfeature score.

**Supplementary Figure 1:** Distribution of PFS for immediate and delayed recall of logical memory. *Note*: PFS = polyfeature score. Vertical dotted line indicates median PFS.

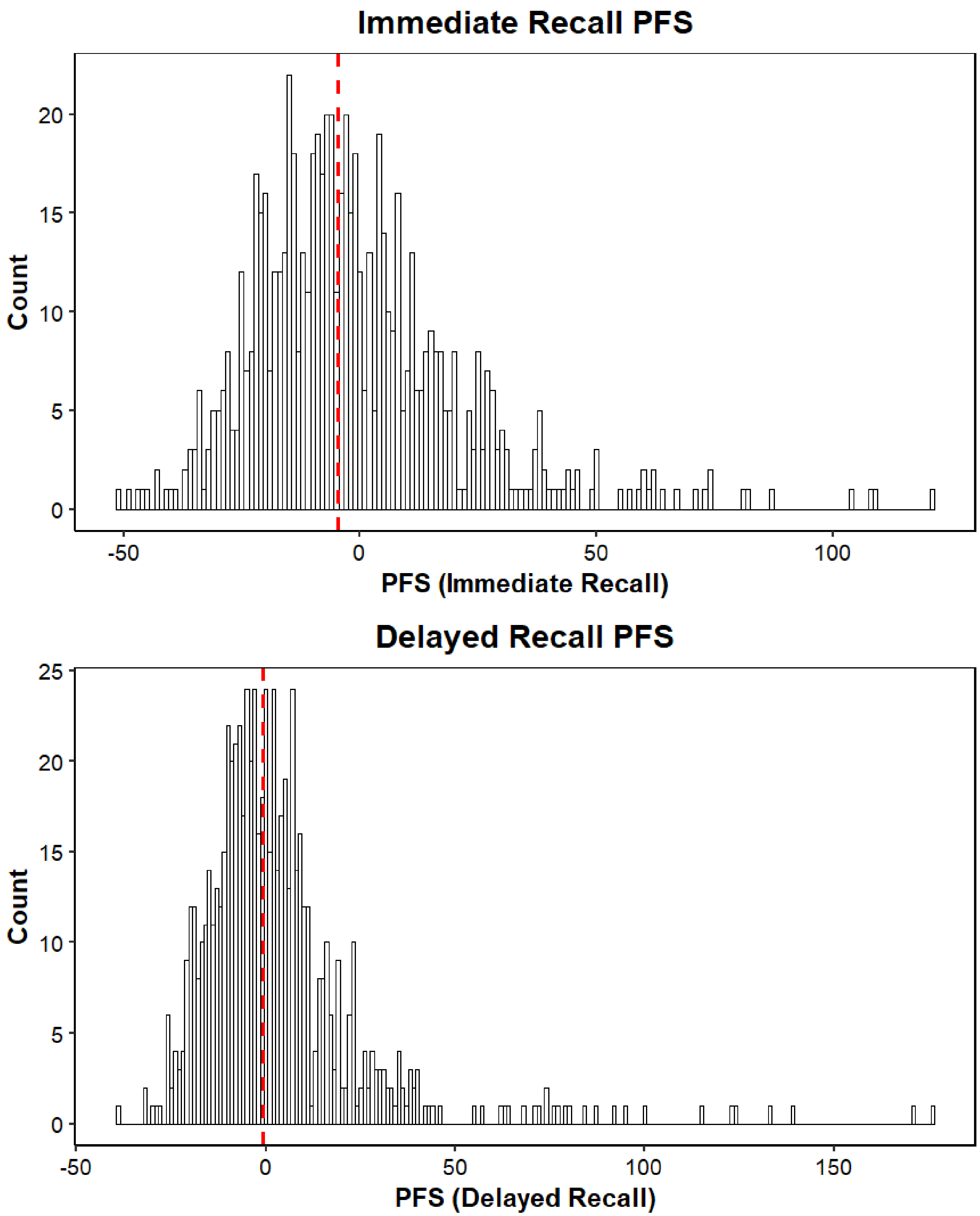

**Supplementary Figure 2**: Relationship between linguistic polyfeature scores (PFS) and traditional logical memory (LM) scores. *Note*: Spearman's rho (ρ) values are reported in the figure.

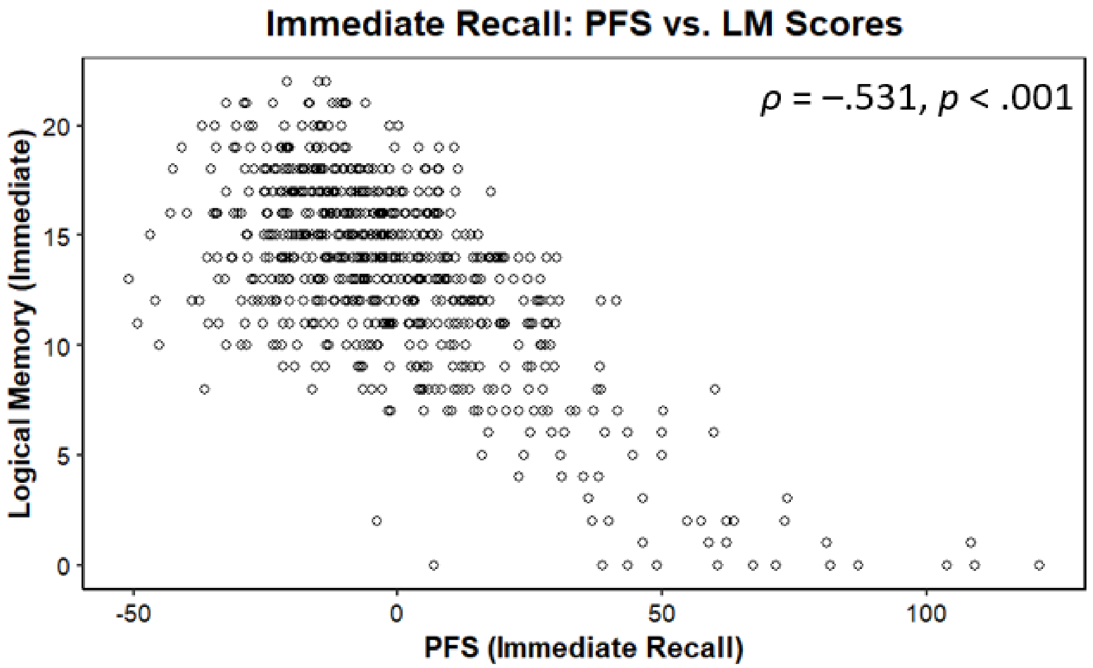


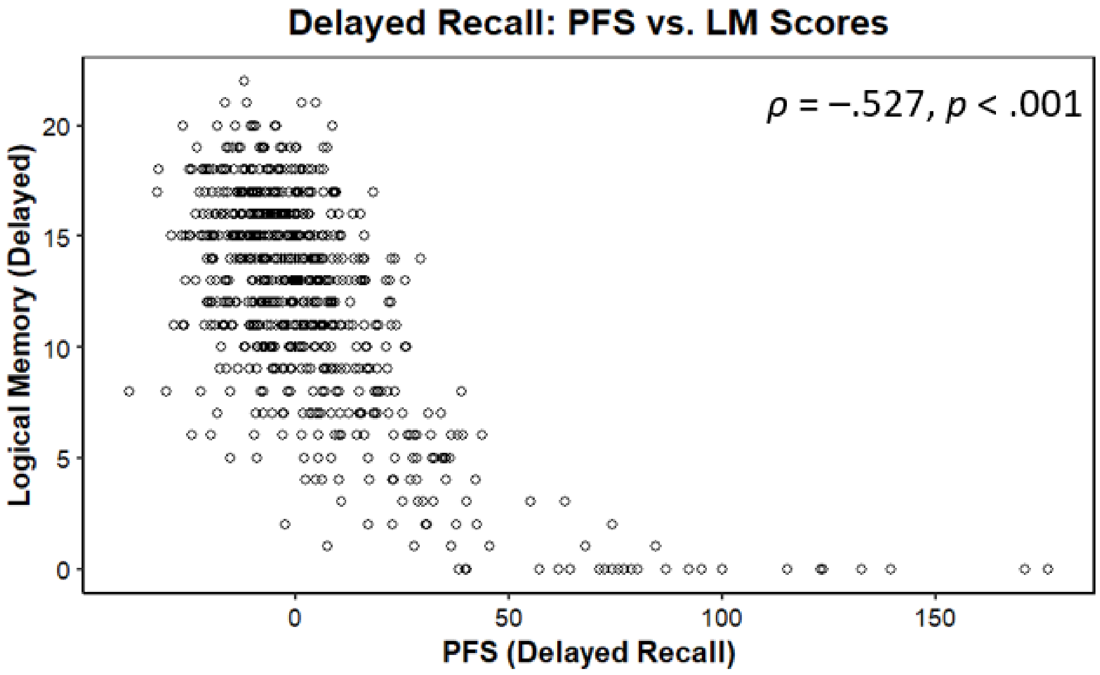

**Supplementary Figure 3**: Delayed recall responses from two participants with traditional logical memory score of 7. *Note*: PFS = polyfeature score.

**A. PFS = 34.2, Consensus review = Cognitively Impaired**

*Okay let's see she had a name. Anna maybe? Oh boy, Anna Thompson? Something like that. I'm not sure of that. Anyway, she um, oh boy... Something in between but she got robbed, and the policemen... ah I guess tried to ask people for help for her and... She had four kids... I can't remember that much of that let's see, there's more stuff in it. You took- you took a good story there... I don't remember now. I know there's more to it but...*

**B. PFS = -18.2, Consensus review = Normal Cognition**

*Really? Ok um, Anna who worked at a school went to the police and told them that she had been robbed. She has three children, and the police were very sympathetic and were gonna try to help her out.*
